# Accelerated biological aging six decades after prenatal famine exposure

**DOI:** 10.1101/2023.11.03.23298046

**Authors:** Mengling Cheng, Dalton Conley, Tom Kuipers, Chihua Li, Calen Ryan, Jazmin Taubert, Shuang Wang, Tian Wang, Jiayi Zhou, Lauren L. Schmitz, Elmar W. Tobi, Bas Heijmans, L.H. Lumey, Daniel W. Belsky

## Abstract

To test the hypothesis that early-life adversity accelerates the pace of biological aging, we analyzed data from the Dutch Hunger Winter Families Study (DHWFS, N=951). DHWFS is a natural-experiment birth-cohort study of survivors of in-utero exposure to famine conditions caused by the German occupation of the Western Netherlands in Winter 1944-5, matched controls, and their siblings. We conducted DNA methylation analysis of blood samples collected when the survivors were aged 58 to quantify biological aging using the DunedinPACE, GrimAge, and PhenoAge epigenetic clocks. Famine survivors had faster DunedinPACE, as compared with controls. This effect was strongest among women. Results were similar for GrimAge, although effect-sizes were smaller. We observed no differences in PhenoAge between survivors and controls. Famine effects were not accounted for by blood-cell composition and were similar for individuals exposed early and later in gestation. Findings suggest in-utero undernutrition may accelerate biological aging in later life.

**Significance Statement:** Environmental conditions during gestation are hypothesized to shape health across the life course. The Dutch Hunger Winter, a famine caused by a German blockade of the Western Netherlands in late 1944 and ended by the allied liberation of the Netherlands in Spring 1945, has been studied as a “natural experiment” in which the timing of a child’s conception determined their exposure to severe under-nutrition during gestation. We applied this natural-experiment design to test effects of in-utero adversity on midlife biological aging, as measured by epigenetic clocks. We found that individuals with in-utero famine exposure had a faster pace of biological aging six decades later. The environmental conditions surrounding pregnancy have potential to shape aging trajectories for the next generation.

## Introduction

Insults to early-life development are predicted by theory to impact trajectories of healthy aging (1–4). Consistent with this hypothesis, longitudinal observational studies have identified associations between early-life conditions and later-life health outcomes (5). But establishing the causality of such associations is difficult due to potential confounding effects of family history and other factors that may affect both early-life development and later aging outcomes (3, 6). Natural experiments, which seek to overcome this obstacle to causal inference, are study designs that take advantage of historical events beyond the control of individuals or their families that impact a subset of otherwise comparable individuals in a population. An established natural-experiment design for investigating effects of early-life adversity on later-life health is in-utero exposure to famine (7). In studies of the Dutch Hunger Winter (1944-5), Siege of Leningrad (1941-4), Holodomor famine in Ukraine (1932-3), and Great Chinese Famine (1959-61), survivors of in-utero famine exposure exhibit higher burdens of multiple aging-related diseases and have shorter lifespans as compared to unexposed individuals born before or after famine or in adjacent, unaffected regions (8–13). Within the Fetal Origins and Developmental Origins of Health and Disease literatures, these observations are often interpreted as reflecting in-utero programming of risk for cardiovascular and metabolic disease later in life (4, 14, 15). However, an alternative hypothesis is that famine-induced insult in early life impairs the development of more general robustness and resilience capacities of the organism, resulting in accelerated systemic decline with aging.

To explore this alternative hypothesis, we analyzed blood DNA methylation (DNAm) data collected in a natural-experiment study of in-utero famine exposure to test differences in the pace and progress of biological aging between famine-survivors and matched controls. The Dutch Hunger Winter Families Study (DHWFS) enrolled a cohort of survivors of in-utero exposure to the Dutch Famine (1944-5), matched controls born before or after the famine in the same hospitals as the survivors, and their same-sex siblings (16). We compared famine survivors to unexposed controls on three DNAm measures of biological aging linked in prior studies with histories of early-life adversity, the DunedinPACE, GrimAge, and PhenoAge DNAm clocks (17–19). Our analysis further explored differences in the effects of famine between women and men and by gestational timing of exposure, and tested consistency of findings in both between-and within-family comparisons.

## Results

We analyzed data for N=951 cohort members with available DNAm data (N=487 famine survivors, N=159 time controls, N=305 sibling controls; **Table 1**). The characteristics of this analysis sample were similar to the Dutch Hunger Winter Families Study interview sample (***SI Appendix*, Table S1**).

**Table 1.** Characteristics of the Dutch Hunger Winter Families Study DNA methylation sample. The table shows characteristics of the analysis sample overall (left column) and the famine-exposed and control groups (middle and right columns).

| **Panel I: DHWFS sample (N=951)** |  |  |  |  |  |  |  |  |
| --- | --- | --- | --- | --- | --- | --- | --- | --- |
|  | **DHWFS** |  | **Famine-exposed** |  | **Controls** |  |  |  |
|  | **(N=951)** |  | **(N=487)** |  | **Time controls**<br>**(N=159)** |  | **Sibling controls**<br>**(N=305)** |  |
|  | Mean/ % | (SD) | Mean/ % | (SD) | Mean/ % | (SD) | Mean/ % | (SD) |
| Age (years) | 58 | (4) | 59 | (1) | 59 | (2) | 57 | (6) |
| Men (%) | 45% |  | 47% |  | 45% |  | 42% |  |
| Duration of exposure (weeks) |  |  | 17 | (7) |  |  |  |  |
| DunedinPACE | 0.97 | (0.11) | 0.97 | (0.11) | 0.95 | (0.11) | 0.97 | (0.11) |
| PC GrimAge | 69.92 | (5.03) | 70.42 | (4.20) | 70.18 | (4.60) | 68.98 | (6.20) |
| PC PhenoAge | 50.18 | (5.87) | 50.61 | (5.18) | 50.61 | (5.10) | 49.27 | (7.07) |

Our analysis proceeded in three steps. First, to test the hypothesis that in-utero famine exposure contributed to accelerated biological aging, we compared DNAm measures of pace of aging (DunedinPACE) and biological age (GrimAge and PhenoAge) between famine-survivors and controls. Second, we conducted dose-response analysis to test if participants who were exposed to the famine for more weeks of gestation exhibited larger famine effects as compared with those exposed for fewer weeks of gestation. Third, to explore specificity of famine effects to exposure during specific periods of gestation, we classified famine survivors according to when in gestation they were exposed, as described in the Methods section, and computed effect estimates for each window of exposure. In each step, we conducted analysis (a) in the full DHWFS; (b) using a between-families comparison of famine-survivors to unexposed time controls born immediately before or after the famine; and (c) using a within-family comparison of famine-survivors to their unexposed same-sex siblings. We also explored whether associations of famine exposure with biological aging differed between men and women.

### In-utero famine exposure was associated with faster biological aging as measured by DunedinPACE

Cohort members exposed to famine in utero had faster pace of aging compared with unexposed cohort members (DunedinPACE β=0.15, 95% CI [0.03, 0.28], *p*=.018). Differences between famine survivors and controls were of smaller magnitude for GrimAge and PhenoAge and not statistically different from zero at the alpha=0.05 level (βs<0.10, *p>*.099). Results were similar in analysis restricted to include only famine survivors and unrelated time controls. Results are shown in **Fig. 1*A*** and reported in ***SI Appendix*, Table S2**; full results for all biological aging measures are reported in ***SI Appendix*, Table S3**; a correlation matrix is shown in ***SI Appendix*, Fig. S1**.

**Fig. 1.**
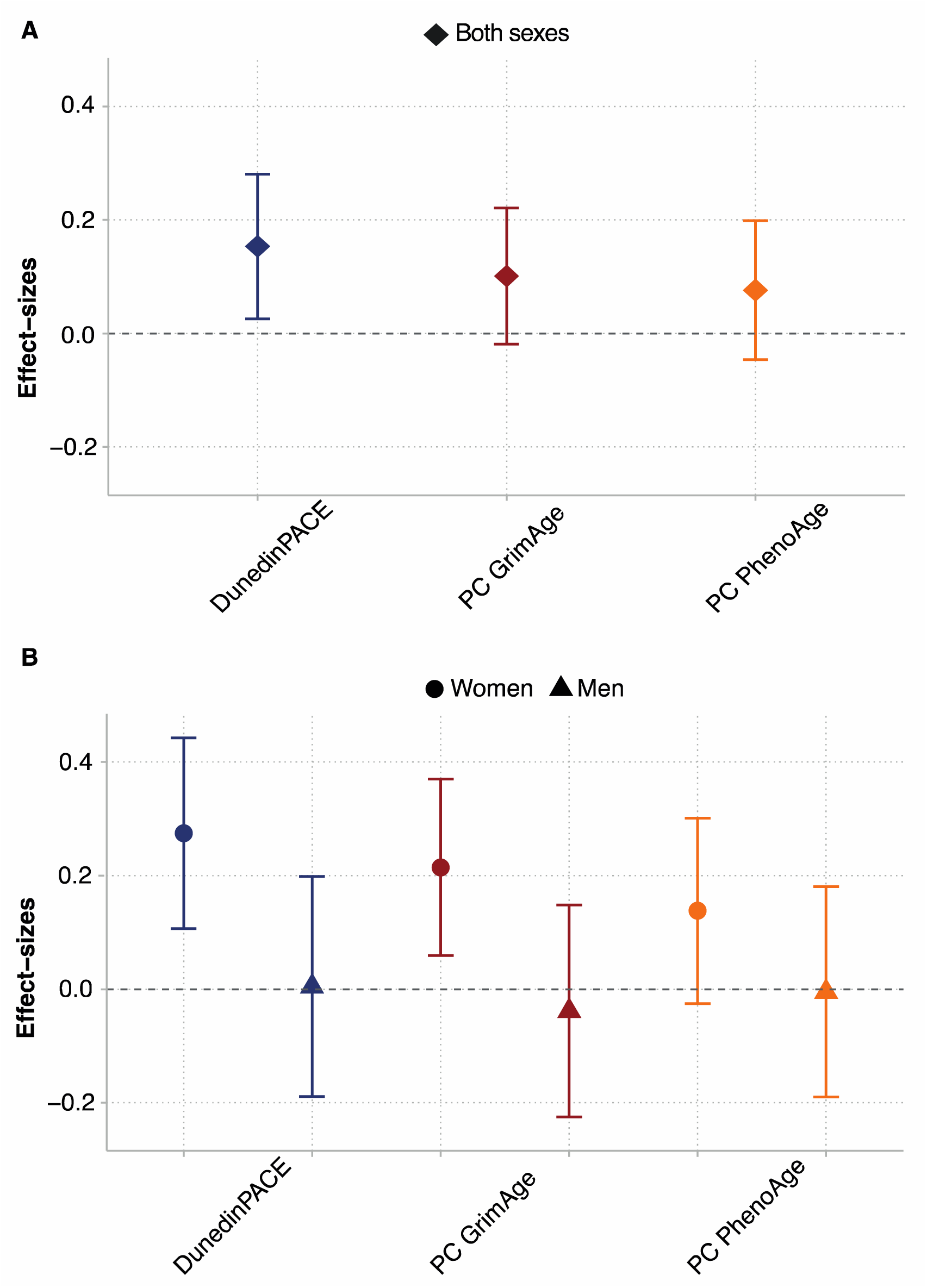
Differences in biological aging between survivors of in-utero famine exposure and unexposed control participants in the Dutch Hunger Winter Families Study. The figure shows effect-sizes of in-utero famine exposure associations with three DNA methylation (DNAm) measures of biological aging, DunedinPACE, PC GrimAge, and PC PhenoAge (N=951). Panel A shows effect-sizes estimated in the full cohort. Panel B shows effect-sizes estimated for women and men separately. Effect-sizes were estimated from generalized estimating equation regressions and are denominated in standard-deviation units of the aging measures, interpretable as Cohen’s d values. Error bars show 95% confidence intervals.

### Longer prenatal famine exposure was associated with faster biological aging as measured by DunedinPACE

In dose-response analysis, cohort members who were exposed to the famine for more weeks of gestation had faster biological aging (per 10-weeks of exposure DunedinPACE β=0.08, 95% CI [0.02, 0.14], *p*=.013). There were no dose response effects for GrimAge and PhenoAge (βs=0.04, *p>*.160). Results are reported in ***SI Appendix*, Table S4**; full results for all biological aging measures are reported in ***SI Appendix*, Table S5**.

### No timing-specificity for famine exposure in predicting biological aging as measured by DunedinPACE

The effects of in-utero famine exposure may vary depending on when in gestation famine exposure occurs. We estimated associations of famine exposure with biological aging at each of six time windows from the preconception period through the end of gestation. Because cohort members could be exposed in multiple time windows, we included indicator variables for exposure in each time window in the same regression. Effect-sizes for DunedinPACE ranged from −0.01 to 0.18 and were somewhat larger for later gestational exposure windows. Effect-sizes for GrimAge ranged from −0.08 to 0.12. Effect-sizes for PhenoAge ranged from −0.19 to 0.28. For GrimAge and PhenoAge, there were no gestational timing patterns in effect-sizes. Effect-sizes are reported in ***SI Appendix*, Table S6**.

### Sex differences in associations of in-utero famine exposure with biological aging

We conducted exploratory analysis of sex differences in famine effects using sex-stratified regressions and analysis of effect-measure modification. In stratified analysis, famine effects were consistently larger for women and were near zero for men (**Fig. 1*B***; ***SI Appendix*, Table S2**). Findings from sex-stratified dose-response analysis showed similar results (***SI Appendix*, Table S4**). In sex-stratified analysis of gestational timing, results were different for women and men (***SI Appendix*, Table S6**). For women, DunedinPACE effect-sizes were similar across gestational time windows (effect-sizes ranged from β=0.07-0.25). In contrast, for men, DunedinPACE effect-sizes were largest for later-gestational exposures and smaller for exposure in early gestation (effect-sizes ranged from β=−0.16-0.19). This pattern was similar for GrimAge. There was no consistent pattern for PhenoAge. Results are shown in **Fig. 2**. Formal tests of effect-measure modification found sex differences were statistically different from zero at the p<0.05 level for DunedinPACE and GrimAge, but not PhenoAge.

**Fig. 2.**
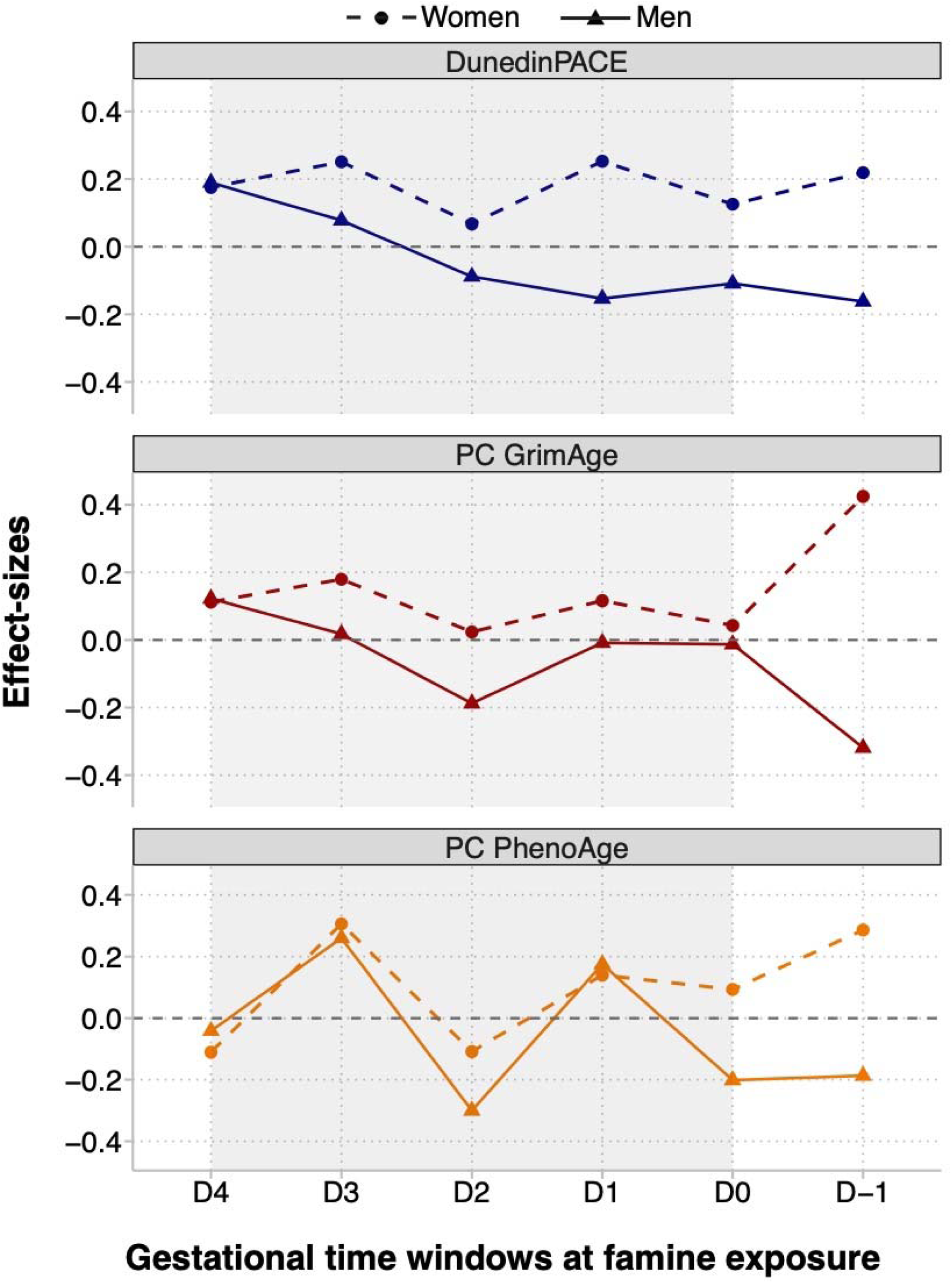
Differences in biological aging between survivors of in-utero famine exposure and unexposed control participants in the Dutch Hunger Winter Families Study by gestational timing of famine exposure. The figure shows effect-sizes estimated for famine exposure during six gestational time windows. Famine-exposed participants were exposed during up to two periods. The developmental periods are ordered in the x-axis in chronological order relative to the famine. The left-most tick shows effect-sizes for late-gestational exposure (defined as exposure for the final 10 weeks of gestation; N=139 exposed). The second tick to the left shows effect-sizes for exposure during the penultimate 10 weeks of gestation (N=146 exposed). The third tick shows effect-sizes for exposure during the second 10 weeks of gestation (N=125 exposed). The fourth tick shows effect-sizes for exposure during the first 10 weeks of gestation (N=74 exposed). The fifth tick shows effect-sizes for early gestational exposure with duration <10 weeks (N=94 exposed). The right-most tick shows effect-sizes for preconceptual exposure, i.e., for exposure during the period preceding conception (N=52 exposed). Numbers exposed do not add up to the total exposed sample because many participants were exposed in two adjacent periods (N=143). Effect-sizes are reported for DunedinPACE, PC GrimAge, and PC PhenoAge. Effect-sizes were estimated from a multivariate regression in which indicator variables for each exposure window were included as predictor variables along with covariates for sex, age, and age-squared. Effect-sizes are denominated in standard-deviation units of the aging measures, interpretable as Cohen’s d values. Full results are reported in *SI Appendix*, Table S6. Effect-sizes are plotted separately for women (circles) and men (triangles). The figure shows a consistent sex-specific pattern in DunedinPACE and PC GrimAge effect-sizes. Women who survived in-utero famine exposure, whether at early or later gestation, tended to have faster pace of biological aging, as measured by DunedinPACE, and older biological age, as measured by PC GrimAge DNAm clock. In contrast, men who survived later-gestational famine exposure tended to have faster pace of biological aging and older biological age; whereas men who survived early-gestational famine exposure tended to have slower pace of biological aging and younger biological age, as measured by DunedinPACE and PC GrimAge DNAm clock, respectively. There was no consistent pattern in PC PhenoAge effect-sizes.

### Sibling-comparison analysis

Finally, we repeated our analysis using a sibling-comparison design. This design holds constant all factors that are shared by siblings in a family. In the context of the famine natural experiment, the sibling comparison design aims to rule out the possibility that differences between exposed and unexposed individuals reflect differences between families in their preferences and/or ability to conceive and carry to term a child under famine conditions. In full-sample sibling comparison analysis (n=227 pairs), famine survivors tended to be aging faster than their unexposed same-sex siblings; however, effect-sizes were attenuated by roughly half as compared with the full-sample analysis and were not statistically different from zero. In sex-stratified analysis, differences between sisters discordant for famine exposure (n=129 pairs) were nearly identical to famine-effect estimates from our original models whereas, among brothers (n=98 pairs), effect estimates were near zero or in the opposite direction of our original analysis. Results are reported in ***SI Appendix*, Table S2 and Table S3**.

### Sensitivity analysis

We repeated our analysis with additional covariates for estimated proportions of white blood cell types. In cell-count-adjusted analysis, effect-sizes were similar for DunedinPACE, GrimAge, and PhenoAge (***SI Appendix*, Table S7-9**).

## Discussion

We analyzed blood DNA methylation data from participants in a natural-experiment study of the Dutch Famine to test the hypothesis that in-utero famine exposure would be associated with accelerated biological aging over six decades of follow-up. We found that survivors of in-utero famine exposure had faster pace of biological aging as measured by the DunedinPACE clock. However, differences in biological age measured by the GrimAge and PhenoAge clocks were smaller and less consistent. We did not observe evidence for a timing specific effect of famine exposure on these measures. These findings were robust to covariate adjustment for cell counts and were similar in sibling-difference analysis, although estimates were less precise.

All three of the DNAm clocks we analyzed show evidence of association with morbidity and mortality in other studies (20, 21). A prior quasi-experimental analysis of early-life economic adversity found evidence of in-utero-exposure effects on later-life biological aging measured by both an earlier version of the DunedinPACE clock and the GrimAge clock (19). However, only DunedinPACE showed consistent evidence of association with in-utero famine exposure in the full DHWFS sample. It could be that DunedinPACE is somewhat more sensitive to preclinical health changes occurring in famine survivors as compared with GrimAge.

DunedinPACE was developed from analysis of the rate of physiological decline in midlife adults (22). It is designed to measure the Pace of Aging phenotype (23), defined as the rate of decline in system integrity. GrimAge, in contrast, was developed from analysis of mortality risk in mid-late life adults (24). It is designed to measure biological age, or the current level of system integrity. These design differences may have consequences for sensitivity in the context of midlife follow-up of in-utero famine exposure. Alternatively, the effect-size differences between DunedinPACE and GrimAge were small and could reflect statistical noise. Follow-up in other studies is needed to clarify the significance of the difference in results for the two measures.

Prior studies suggest that exposure during the early phase of gestation may be more impactful (11, 25–27). Our analysis of gestational timing of famine exposure did not find evidence for earlier gestation as a sensitive period. Overall, results suggest that any in-utero exposure is associated with a faster pace of biological aging six decades later.

Our analysis of sex differences in famine effects found larger effects of in-utero famine exposure on DNAm measures of biological aging in women as compared with men. This was observed for all three epigenetic clocks, but was most pronounced for the DunedinPACE and GrimAge clocks. In non-human animals, there is evidence that males are more vulnerable to early-life insults (28). However, prior studies of in-utero famine exposure often reported larger famine effects on cardiovascular and metabolic diseases among women as compared with men (29–32). There is some evidence that selective fertility and/or fetal loss result in fewer male births during periods of famine (33). A result could be that the subset of male babies born are especially robust. This would be consistent with our results. But a reduction in male births is not documented in the case of the Dutch Famine (34, 35). There are not yet analyses of sex differences in mortality among survivors of in-utero exposure to the Dutch Famine; the most comprehensive study of mortality relied on data from military conscripts, who were all men (25). New models in population science suggest that environmental conditions, such as in-utero exposure to famine, can induce substantial variation in sex differences in survival (36). Replication of the observed sex difference in famine effects is needed.

We acknowledge limitations. There is no gold standard to measure biological aging (37). We analyzed the DNAm measures of aging with the best available evidence for reliability and validity. As new measures are introduced, follow-up will be needed. However, agreement between different biological aging measures build confidence that our findings do capture aging processes. We were unable to conduct dose-response analysis of famine-exposure severity. Because of the lack of family-level nutrition data and the consistency of rations across the affected areas of the Netherlands, analysis of exposure severity will need to be conducted in different settings, such as where famine severity was graded across geographic locations or time (9). Survival bias could affect the population of famine survivors alive or in sufficiently good health to be surveyed at follow-up. However, characteristics at birth of the DHWFS participants are similar to those of famine-affected births identified in hospital records but not successfully enrolled in the cohort, including birth weight, length, placental weight, maternal age, and birth order (16). Excess deaths among survivors of in-utero famine exposure by the time of our study were <10% (13). Therefore, any bias is likely to be modest. Moreover, it is expected that healthy-participants and survival biases would bias effect-estimates toward the null because non-participation due to ill health and death would remove the most affected from the population. Therefore, our estimates of famine effects are expected to be conservative. Finally, because the cohort so far lacks follow-up to determine survival differences between famine-exposed and control participants, the extent to which differences observed in measures of biological aging will translate into differences in healthspan and lifespan remains to be determined.

Within the context of these limitations, our findings provide evidence for long-term impacts of in-utero famine exposure that may extend to a wide range of aging-related health outcomes. Now that survivors of in-utero famine exposure are approaching their ninth decade of life, further study of famine births in administrative record data are needed to clarify the scope of famine effects on healthspan and lifespan.

## Materials and Methods

### Study setting: The Dutch Hunger Winter of 1944-1945

The Dutch famine was initiated by a food supply embargo imposed by the German occupying forces in early October 1944. The severity and widespread nature of the famine are well documented (16, 38, 39). Prior to the embargo, nutrition in the Netherlands had generally been adequate. Official rations, which eventually consisted of little more than bread and potatoes, fell below 900 kcal/day in late November 1944, and were as low as 500 kcal/day by April 1945. The macronutrient composition of the ration remained relatively stable over this period, but the composition of non-ration foods changed, with a reduction in the intake of fat. The famine ceased with liberation in May 1945, after which Allied food supplies were distributed.

### Participants

*Famine-Exposed* individuals were identified from review of archival obstetric records. We selected all the 2,417 singleton births between 1 February 1945 and 31 March 1946 at three institutions in famine-exposed cities in the western Netherlands whose mothers were exposed to the famine during or immediately preceding that pregnancy.

*Time Controls* were selected from births at the same hospitals and in the same months of the year as the famine-exposed group during 1943 and 1947 (two years before and two years after the famine). We sampled an equal number of births in each month, allocated across the three institutions according to their size, to obtain 890 singleton births.

Of the total famine-exposed and time-control births, current addresses were able to be traced for 70%. These individuals were invited by mail to join the study and were additionally asked if a *same-sex sibling* born before or after the famine would be available to participate. A total of N=547 of the famine-exposed group, N=176 of the time-control group, and N=308 same-sex unexposed siblings consented to participate and underwent a computer-assisted structured interview by telephone.

Data Collection was conducted in 2003-2005, approximately six decades after the famine. Of the N=1,031 participants who were interviewed, N=971 also participated in a clinic exam (**Fig. 3**). Following the Helsinki guidelines, we obtained ethical approval both from the Institutional Review Board of Columbia University Medical Center and from the Medical Ethical Committee of the Leiden University Medical Center (LUMC). The study participants provided verbal consent in a telephone interview, and in case of clinical examinations, a written informed consent was obtained and additional METC approval for epigenetic studies was later confirmed by the METC of the LUMC.

**Fig. 3.**
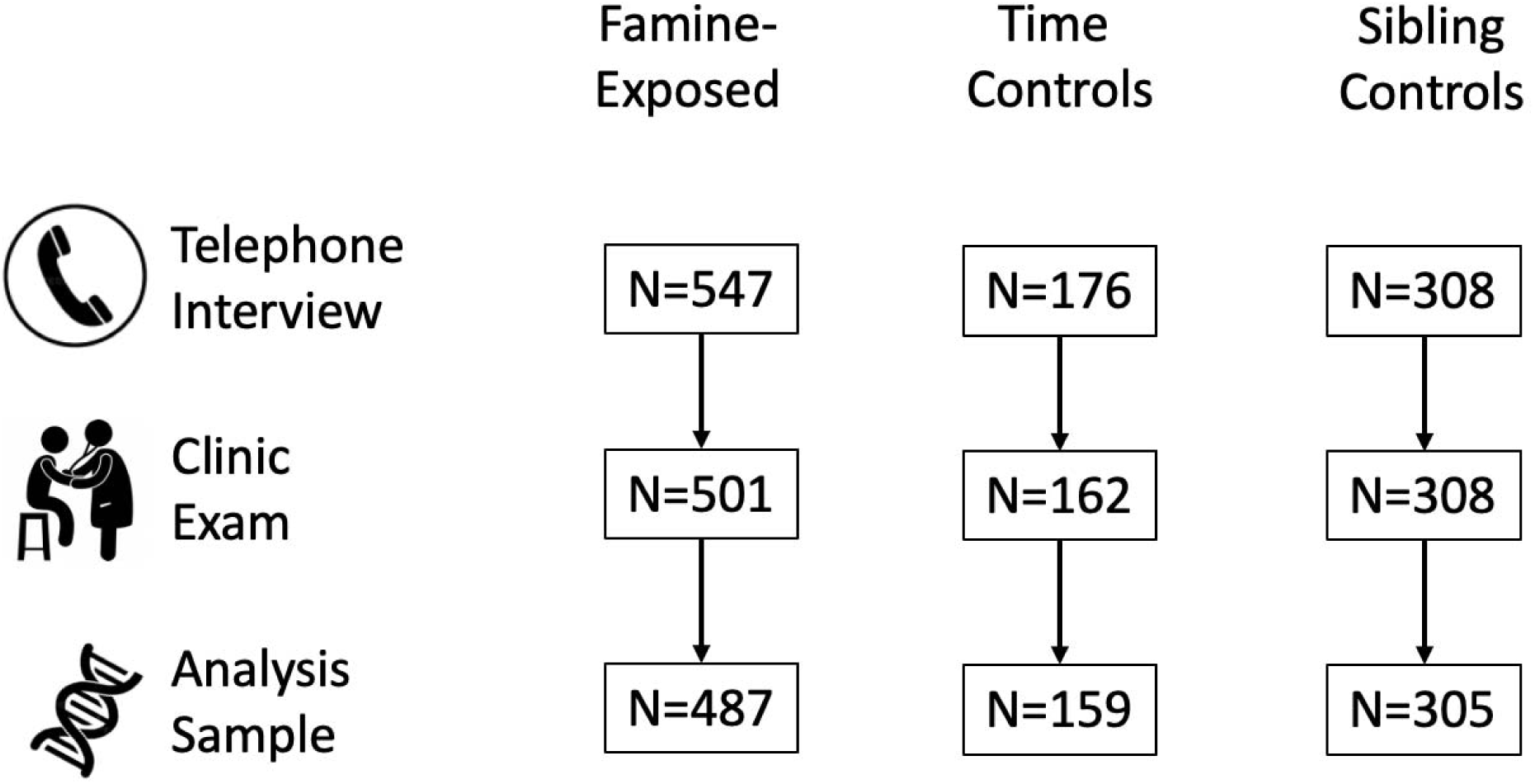
Flow diagram of the Dutch Hunger Winter Families Study. The figure shows how the analysis sample size was arrived at in each step for survivors of in-utero famine exposure, time controls, and same-sex sibling controls. N=1,031 participants completed telephone interviews. Of this group, 971 participated in the clinic exam. DNA extracted from blood samples was analyzed to determine DNA methylation and data passed quality controls for N=951 individuals. The figure illustrates the number of individuals in each exposure and control group included in the telephone interview, clinic examination, and analysis sample.

### Measures

#### Famine exposure

We defined the period of famine from archival records of weekly ration distributions, as described previously (16). Briefly, the start of the famine-exposure period was defined as November 26, 1944, based on the threshold <900 kcal/day of distributed food rations. The end of the famine-exposure period was defined as May 12, 1945, one week following the German surrender. Participants’ exposure during gestation was determined from the date of their mother’s last menstrual period (LMP) and their date of birth. In cases where the LMP date was missing or implausible (12% of births), LMP was estimated from birth-record data on birth weight and date of birth using tables of gender, parity, and birth weight specific gestational ages from the combined birth records of the Amsterdam Midwives School (1948-57) and the University of Amsterdam Wilhelmina Gasthuis Hospital (1931-65).

Famine-exposed participants experienced an average of 17 weeks of gestation during which ration distributions were <900kcal/day. Following prior work with the cohort (29, 40), participants were classified as famine exposed during each of four 10-week gestational periods on the basis of ration distributions. For each individual, average rations were calculated for each 10-week period of gestation and periods with average rations <900 kcal/day were classified as famine-exposed. Among the N=547 participants recruited as famine exposed, N=403 met criteria for exposure in at least one 10-week gestational period. A further N=82 had LMP dates prior to the end of the famine, but fewer than 10 weeks of gestational exposure to ration distributions <900kcal/day. A final N=62 had LMP dates after the end of the famine. Gestational periods for famine-exposed participants and time-controls are shown in **Fig. 4**.

**Fig. 4.**
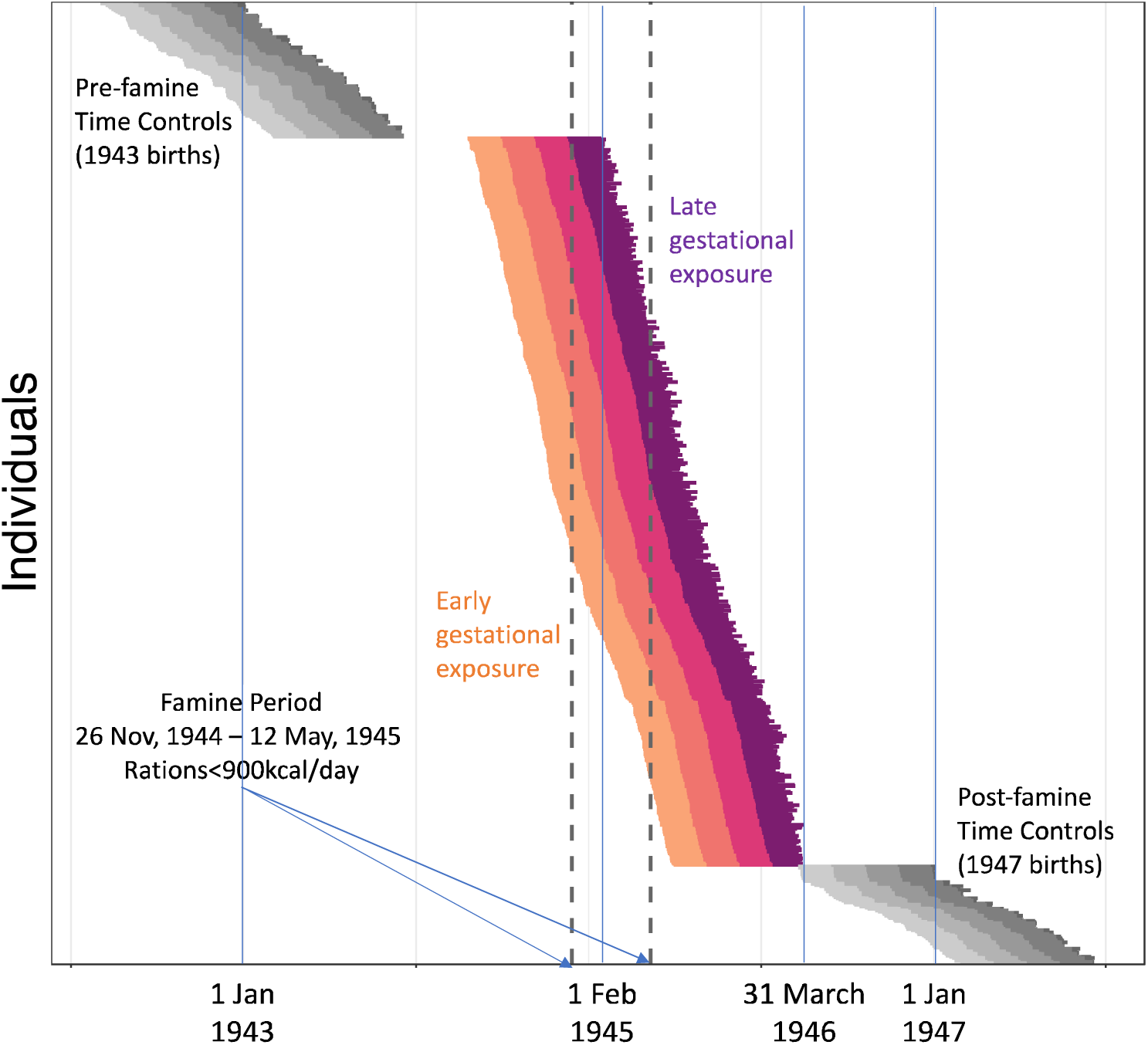
Gestational timing of exposure to famine in the Dutch Hunger Winter Families Study. The figure shows individual gestations of N=547 famine-exposed participants (colored lines) and N=176 time controls (gray lines). Each gestation is plotted as a single horizontal line. The start of the line is the date of the mother’s last menstrual period (LMP). The end of the line is the participant’s date of birth. Individual gestations are plotted from the top of the graph to the bottom, ordered by LMP date. For the famine-exposed participants, the segment of each line showing the first 10 weeks of gestation is colored gold. The segment showing the second 10 weeks is colored orange. The segment showing the third 10 weeks is colored red. The segment showing the last 10 weeks is colored purple. For the time controls, 10-week gestational periods are colored in gray, with lighter shades for the earlier gestational periods. The x-axis shows the date. The vertical dashed lines show the start and end of the famine exposure period (November 26, 1944 - May 12, 1945).

#### DNA methylation measures of biological aging

Biological aging is the progressive loss of system integrity with advancing chronological age (41). Biological aging is thought to arise from an accumulation of cellular-level changes that progressively undermine the robustness and resilience capacity of cells, tissues, and organ systems (42–44). While there is no gold-standard measure of biological aging in humans (37), the current-state of the art are algorithms that combine information from dozens or hundreds of DNA methylation (DNAm) marks, chemical tags on the DNA sequence that regulate gene expression and are known to change with aging (45). These algorithms are often referred to as “epigenetic clocks” (46). We measured biological aging using epigenetic clocks computed from the existing DNA methylation database for the DHWFS.

Briefly, DNA methylation (DNAm) was measured from blood collected at the clinic exam using the Illumina Infinium Human Methylation 450k BeadChip and preprocessed as previously described (26). Further details are provided in the ***SI Appendix*, Supplementary Methods**.

Our primary analysis focused on three epigenetic clocks for which validation data across multiple studies establish robust associations with healthspan and lifespan and sensitivity to exposures known to hasten aging-related health decline: the DunedinPACE clock, which measures pace of aging, and the GrimAge and PhenoAge clocks, which measure biological age. We calculated DunedinPACE using the R code available on GitHub (https://github.com/danbelsky/DunedinPACE). We calculated high-technical-reliability “PC” versions of the GrimAge and PhenoAge clocks developed by the Levine Lab (47) using the code available from GitHub (https://github.com/MorganLevineLab/PC-Clocks). The clocks are described in detail in the ***SI Appendix*, Supplementary Methods**.

There are many other epigenetic clocks, although none with comparable evidence of validity to DunedinPACE, GrimAge, and PhenoAge. Most other clocks were developed to predict differences between individuals in their chronological age (sometimes referred to as “first-generation clocks”). For comparison purposes, we report results in the***SI Appendix*, Table S3 and Table S5** for three of the best-known first-generation clocks, the Horvath, Hannum, and Skin & Blood clocks (46, 48, 49). We also report results for the original versions of three second-generation clocks, the Zhang, GrimAge, and PhenoAge clocks (24, 50, 51). Original versions of the clocks were computed using the methylclock R package (52) and Python code to calculate GrimAge provided by Ake Lu.

### Analysis

The analysis sample for this study was formed from participants in the clinic examination who provided a blood sample from which DNA was extracted and stored at LUMC. For our analysis, DNA were available for N=960 individuals. After quality controls, DNA methylation datasets were available for N=951. These individuals formed our analysis sample.

We used regression analysis to test associations between in-utero famine exposure and DNAm measures of biological aging. First- and second-generation epigenetic clock values have high correlations with chronological age. For analysis and interpretation, the standard approach is to regress clock values on participants’ chronological age values and predict residual values. These values, often referred to as “age acceleration residuals”, aim to quantify the difference between how much aging a person has actually experienced relative to the expectation based on their chronological age. No residualization was performed for DunedinPACE, which is a rate measure and shows only moderate correlation with chronological age. To account for the non-independence of measurements taken from siblings, we used generalized estimating equation (GEE) regressions (53). Our models included covariates for participants’ sex, age, and age-squared at the time of the clinic exam. We explored sex differences in famine effects by repeating analysis with inclusion of product term testing interaction between famine exposure and sex. We repeated our analysis with a control group restricted to the “time controls” born immediately before or after the famine. We tested consistency of results in within-family comparisons of siblings using sibling-fixed-effects (FE) regressions (54). We tested the sensitivity of associations between famine exposure and biological aging to differences between participants in leukocyte composition of DNA samples by repeating analysis with additional covariates for DNAm estimates of leukocyte proportions estimated using the Houseman equations (55).

## Funding sources

This research was supported by National Institute on Aging grants (R01AG066887; R01AG042190). DWB is a fellow of the Canadian Institute for Advanced Research Child Brain Development Network. LHL was supported by a NIDI/NIAS/UMCG Fellowship, Royal Netherlands Academy of Sciences KNAW at the Netherlands Institute for Advanced Study. MC was supported by the Swiss National Centre of Competence in Research “LIVES - Overcoming vulnerability: Life course perspectives” financed by the Swiss National Science Foundation (51NF40-185901) and the European Union Horizon 2020 Research and Innovation Programme under the Marie Sklodowska-Curie Grant (801076). LLS was supported by National Institute on Aging grant (R00AG056599). EWT was supported by a VENI grant from the Netherlands Organization for Scientific Research (91617128) and by the Joint Programming Initiative (JPI HDHL proposal number 655) via ZonMw in The Netherlands (529051023).The funders had no role in study design, data collection and analysis, decision to publish, or preparation of the manuscript.

## Data Availability

The Dutch Hunger Winter Families Study (DHWFS) is registered as part of the LUMC Biobank Ageing (B15.027). To protect the privacy of research participants, DHWFS data are available to access through the Leiden University Medical Center Biobank from within the secure Leiden University Medical Center network environment. Approval of access by the DHWFS investigators is required. Requests can be made to corresponding author Daniel W Belsky or to the other Study PIs, Lambert H. Lumey (Columbia University Mailman School of Public Health) and Bastiaan T. Heijmans (Leiden University Medical Center).

## Notes

**Conflict of Interest** DWB is listed as an inventor on the Duke University and University of Otago Invention DunedinPACE, which is licensed to TruDiagnostic.

### Competing Interest Statement

DWB is listed as an inventor on the Duke University and University of Otago Invention DunedinPACE, which is licensed to TruDiagnostic.

### Author Declarations

Institutional Review Board of Columbia University Medical Center and Medical Ethical Committee of the Leiden University Medical Center gave ethical approval for this work.

